# The Potential Effect of Ending CDC Funding for HIV Tests: A Modeling Study in 18 States

**DOI:** 10.1101/2025.09.19.25336182

**Authors:** Ruchita Balasubramanian, Melissa Schnure, Ryan Forster, William P. Hanage, D. Scott Batey, Keri N. Althoff, Kelly A. Gebo, David W. Dowdy, Maunank Shah, Parastu Kasaie, Anthony T. Fojo

## Abstract

**Background:** Timely HIV diagnosis and treatment is critical to preventing transmission. The US Centers for Disease Control and Prevention (CDC) provides funding for HIV testing to local health departments and community organizations. We sought to estimate the number of additional HIV infections that would result from ending or interrupting CDC funding for HIV tests in US states.

**Methods:** We used a validated model of HIV transmission to simulate HIV epidemics in 18 US states. We projected incidence forward under three scenarios where all CDC-funded HIV testing ends in October 2025 and (1) never resumes, (2) returns to previous levels between January and December 2027, and (3) returns from January to December 2029. We calculated the excess incident HIV infections compared to a scenario where CDC-funded testing continues uninterrupted.

**Results:** If CDC funding for HIV tests were to end on October 1, 2025, we project 12,719 additional HIV infections across 18 states by 2030 (95% Credible Interval 4,547 to 21,896) – an increase of 10%. The projected effects varied by state, ranging from a 2.7% increase in Washington (1.0 to 4.7%) to a 29.9 increase in Louisiana (9.4 to 59.9%). States that perform more CDC-funded tests and states with more rural HIV epidemics were projected to see greater rises in incidence.

**Conclusions:** Disruptions to CDC-funded HIV testing would substantially increase new infections, particularly in states with more rural epidemics. These findings demonstrate the value of the CDC’s HIV testing activities in curbing the spread of HIV in the US.

**Summary:** A complete cessation of CDC-funded HIV testing could cause a 10% increase in infections by 2030 across 18 U.S. states, with this impact disproportionately affecting rural states. Mobilizing access to alternative HIV testing would be critical to mitigating this impact.

## Introduction

HIV imposes a substantial health burden in the US, with over one million persons living with HIV as of 2023^1^. Timely diagnosis and treatment of HIV is critical to preventing transmission^1,2–4^. People with HIV (PWH) who are virally suppressed on antiretroviral therapy are non-infectious. Even before treatment, PWH reduce behaviors associated with transmission once they are aware of their status^5,6^.

The US Centers for Disease Control and Prevention (CDC) financially supports the bulk of HIV testing by state and local health departments, as well as supporting testing by community-based organizations^7^. In 2021, CDC funding supported 1,736,850 tests, resulting in 8,149 new HIV diagnoses^8^. While CDC’s testing data are not directly comparable to its surveillance data, these diagnoses correspond to over 20% of the 35,763 total diagnoses recorded in the US in 2021. In some states, this ratio is much higher, approaching 50% (or more) of diagnoses in South Carolina, Alabama, and Tennessee^1,8^.

In general, HIV testing is an efficient means of HIV prevention. CDC-funded tests are particularly efficient and are disproportionately used in demographic subgroups with high rates of HIV infection^7^. These CDC-funded HIV testing activities, however, may be subject to funding cuts in the future^9^.

Disruptions to CDC-funded HIV testing could have a significant impact on HIV incidence in the US^10^. Mathematical models can be a useful tool to help forecast the impacts of HIV health policy^11,12^. We used a validated model of HIV transmission in the US to project the potential impact of the cessation or interruption of CDC-funded HIV testing on state-level HIV-epidemics.

## Methods

### Model Structure

The Johns Hopkins Epidemiologic and Economic Model (JHEEM) is a validated, dynamic, compartmental model of HIV transmission that has been calibrated to cities and states in the US and is stratified by age, sex, race/ethnicity, and risk-status for HIV acquisition^11,13^.

To represent the impact of CDC-funded HIV testing, we expanded the JHEEM to simulate, for each demographic stratum, (1) the proportion of HIV tests that are funded by the CDC and (2) the proportion of new diagnoses that are made by CDC-funded HIV tests. These proportions were each modeled using a logistic equation, parameterized with terms for age, race, sex, risk factor, and time (see Supplement).

### Study Setting

We simulated HIV epidemics in 18 states: Alabama, Arizona, California, Florida, Georgia, Illinois, Kentucky, Louisiana, Maryland, Mississippi, Missouri, New York, Ohio South Carolina, Tennessee, Texas, Washington, and Wisconsin. These states were chosen for geographic distribution and balance of urban/rural composition, mix of Medicaid expansion status, and prioritization in the *Ending the HIV Epidemic* initiative^14^.

### Model Calibration

JHEEM’s calibration has been described previously; briefly, we ran an Adaptive Metropolis Sampler for 1,000,000 iterations in each state and retained a set of 1,000 well-fitting simulations^11^. These simulations reproduce local epidemiological measures of the epidemic, including new diagnoses, prevalent cases, and proportion of the general population who report being tested for HIV during the preceding year.

For each of the 1,000 base JHEEM simulations per state, we ran another Adaptive Metropolis Sampler for 300 iterations to calibrate the parameters for the logistic models for the proportion of HIV tests funded by CDC and the positivity among CDC-funded HIV tests. We derived prior distributions based on national data and formulated a likelihood for two calibration targets drawn from annual CDC HIV Testing reports from 2011 to 2019: (1) the number of CDC-funded tests in each state and (2) the rate of positivity (excluding known cases) among CDC-funded tests (see Supplement)^15^.

### Modeled Scenarios

In addition to a scenario where CDC funding for HIV tests continues uninterrupted, we simulated three scenarios (Supplement Figure S1): (1) “Cessation” - CDC funding for HIV tests stops on October 1, 2025, and CDC-funded tests linearly decline to zero by December 31, 2025; (2) “Brief Interruption” - CDC-funded testing ends as in “Cessation”, but then resumes in 2027, rising linearly from zero on December 31, 2026, to previous daily volumes by December 31, 2027. (3) “Prolonged Interruption” - CDC-funded testing ends as in “Cessation”, but then resumes in 2029, rising linearly from zero on December 31, 2028, to previous daily volumes by December 31, 2029. (Supplement Figure S1)

In the absence of CDC funding, presumably some individuals who would otherwise have received a CDC-funded HIV test would get tested by other means, such as private insurance or in an emergency department. However, this proportion is not well characterized: no studies to our knowledge have examined the withdrawal of public funding for HIV tests. One evaluation of the roll-out of a CDC-funded program to distribute HIV self-tests indicated that, of the 206,637 survey respondents who took one or more CDC-funded self-tests, 52% had not been tested in the past year^16^.

To quantify this uncertainty, each of the 1,000 simulations per state sampled a different value of a parameter representing the proportion of CDC-funded tests that would be replaced through other means. We sampled these values from a beta distribution with a mean of 50% based on results from the CDC’s self-test program and a wide 95% confidence interval of 20% to 80%.

In all scenarios, we projected the HIV epidemic in each state to 2030, assuming continuation of current trends in transmission, suppression, uptake of pre-exposure prophylaxis (PrEP), and testing not funded by the CDC, with randomly sampled variation^11^.

### Outcomes

Our primary outcome was the projected relative excess incident HIV infections from 2025 to 2030:

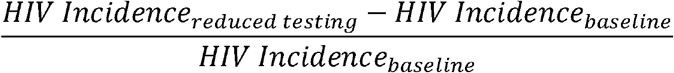

where the “baseline” scenario assumes continued CDC-funded testing at its current trajectory. Secondary outcomes included (a) the absolute number of excess HIV infections from 2025 to 2030, (b) the absolute and relative number of infections through 2035, and (c) the number of CDC-funded tests foregone per excess infection from 2025 to 2030. For each outcome in each state, we calculated the mean across 1,000 simulations and the 95% credible interval (CrI), defined as the 2.5th and 97.5th percentiles of those simulation results.

### Secondary Analyses

To evaluate potential determinants of state-level variation, we calculated Spearman correlation coefficients between our primary outcome (relative projected excess incident HIV infections from 2025 to 2030) and four factors (averaged for each state across simulations): (1) the proportion of HIV tests in 2025 that the CDC funded; (2) the proportion of HIV diagnoses in 2025 that were made with CDC-funded tests; (3) the transmission rate of HIV in 2025; and (4) the “urbanicity” of each state’s HIV epidemic in 2021, defined as the mean across counties (weighted by each county’s fraction of statewide HIV prevalence) of the proportion of people living in urban areas (per the 2020 census)^17^. Because we modeled only 18 states, we calculated a univariate correlation with each determinant separately. We visualized these relationships using scatterplots.

### Sensitivity Analyses

To assess the sensitivity of our primary outcome to influential parameters in each state, we calculated partial rank correlation coefficients, across the 1,000 simulations in each state, for parameters that governed either (a) the proportion of HIV tests funded by the CDC, (b) the proportion of diagnoses made with CDC-funded tests, or (c) the proportion of CDC-funded tests that would be obtained by other means if funding ends. We assessed the impact of each parameter by calculating the primary outcome among the 200 simulations with the highest values of each parameter vs. the 200 simulations with the lowest values (see Supplement)^18^.

### Web Tool

All simulations are available through our interactive web tool at www.jheem.org/cdc-testing.

## Results

Our simulations closely matched the number of CDC-funded HIV tests and positivity rate by state (Supplement Figures S2-7 and online at www.jheem.org/cdc-testing). If CDC funding for tests continues uninterrupted, our model projects 128,900 incident infections from 2025 to 2030 across all 18 states (95% CrI 123,565 to 135,535), and 222,706 infections by 2035 (95% CrI 210,324 to 237,189).

If CDC-funded testing ends permanently in 2025 (“Cessation”), we project 12,719 excess HIV infections by 2030 across the 18 states (95% CrI: 4,547 to 21,896) - an increase of 9.9% (95% CrI 3.6 to 16.9%) compared to continuation of current testing volume, despite plausible levels of replacement tests through other sources of testing. This negative impact varied substantially by state, ranging from a 2.7% increase in HIV infections in Washington state (95% CrI 1.0 to 4.7%) to a 29.9% increase in Louisiana (95% CrI 9.4 to 59.9%) - illustrated in Figures 1, 2, and 3 and online at www.jheem.org/cdc-testing. If CDC-funded testing does not resume, we project 33,691 excess infections by 2035 (95% CrI 11,327 to 60,161) – an increase of 15.1% (95% CrI 5.2 to 26.9% - see Supplement Figure S8).

**Figure 1.**
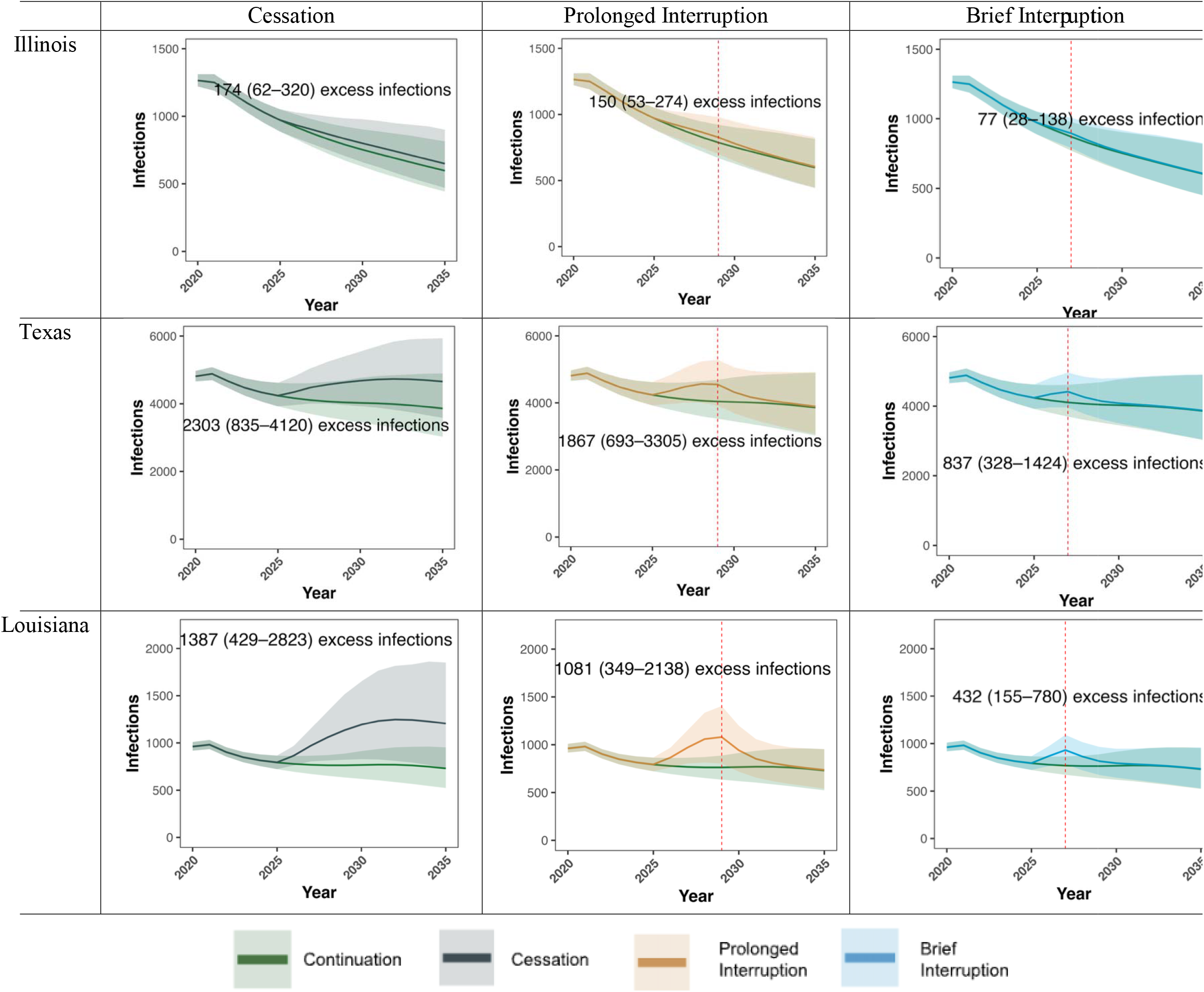
Projected HIV Infections in Illinois, Texas and Louisiana if CDC-funded HIV Testing is Disrupted

**Figure 2.**
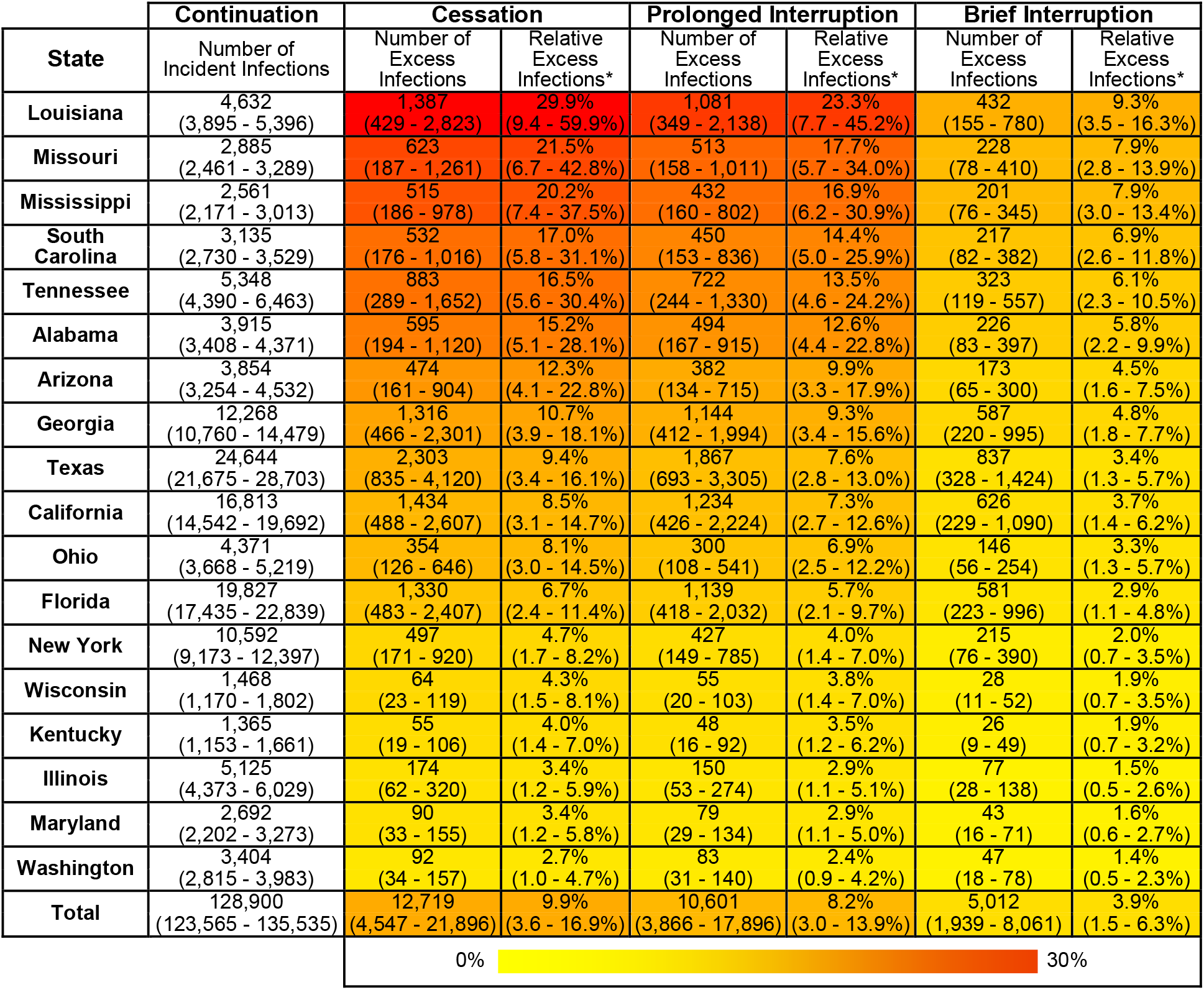
Projected Excess HIV Infections if CDC-funded HIV Testing is Disrupted

**Figure 3.**
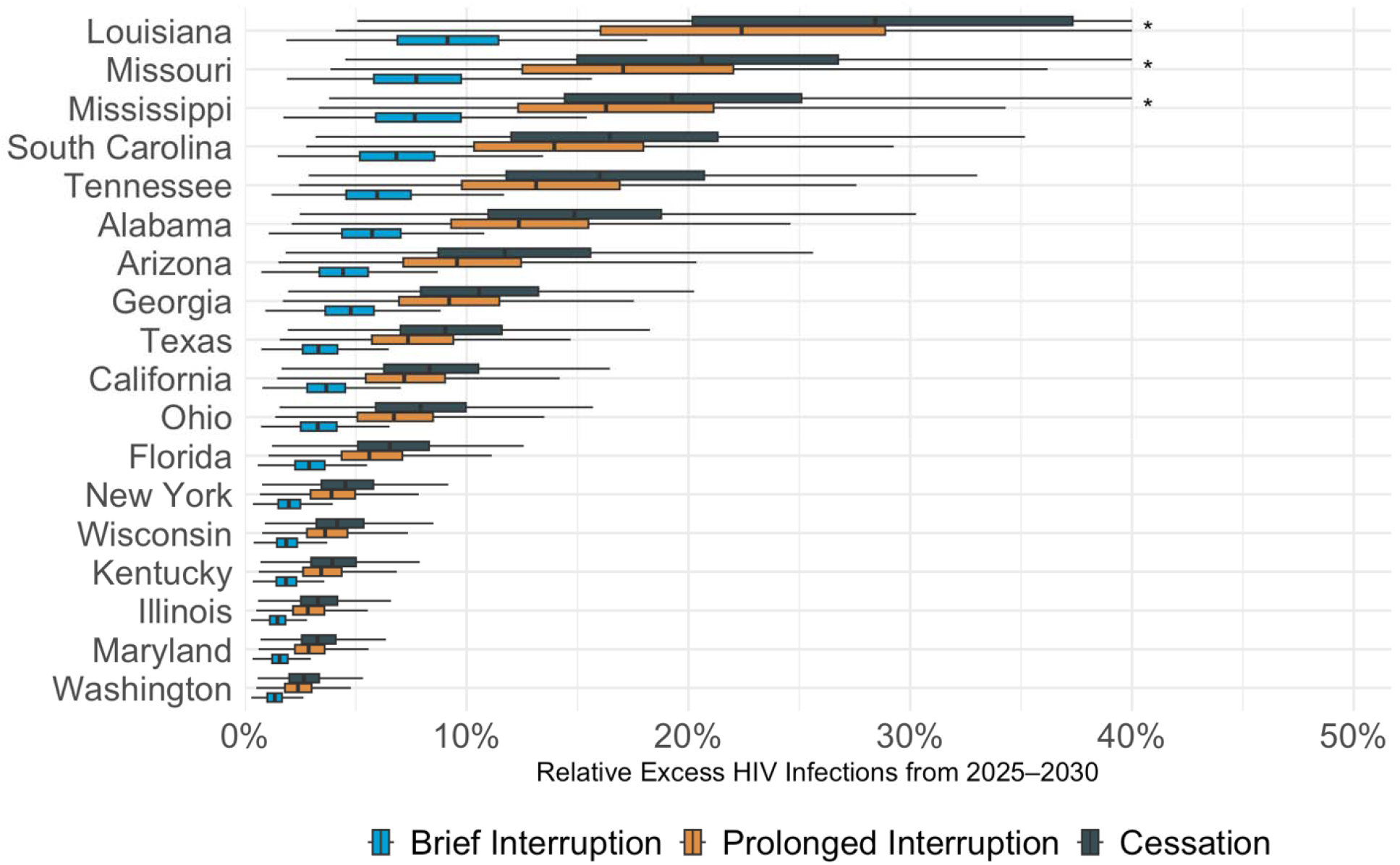
Projected Excess HIV Infections if CDC-funded HIV Testing is Disrupted

If CDC funding for tests were to be restored in 2029 and testing returns to current levels by the end of that year (“Prolonged Interruption”), our model projects 10,601 (95% CrI 3,866 - 17,896) excess HIV infections by 2030, an increase of 8.2% (95% CrI 3.0 to 13.9%) across the 18 selected states. By 2035, we project 6.4% more infections (95% CrI 2.4 to 10.7%) than if testing had continued uninterrupted. If CDC-funded testing returns to baseline levels by the end of 2027 (“Brief Interruption”), the model projects 5,012 (95% CrI 1,939 to 8,061) excess infections by 2030, an increase of 3.9% (95% CrI 1.5 to 6.3%). By 2035, we projected new infections to be 2.8% higher (95% CrI 1.1 to 4.5%).

We project that the increases in infections would accrue more in young adults, with an estimated 13% (95% CrI 5 to 22%) increase in incidence among 13-34-year-olds by 2030 versus 6% (95% CrI 2 to 11%) among those over age 35. Excess infections are also projected to be higher among men who have sex with men (11% increase, 95% CrI 4 to 20%) and heterosexual men (9% increase, 95% CrI 3 to 16%) than among women (7%, 95% CrI 3 to 12%). We do not project significant differences by race, with an expected 11% increase in infections (95% CrI 4 to 19%) among Black adults, 9% (95% CrI 3 to 15%) among Hispanic adults, and 9% 95% CrI (3 to 16%) for non-Black, non-Hispanic adults.

Among all parameters in the model, the proportion of diagnoses made by CDC-funded tests at the state level and the proportion of the state’s HIV tests that the CDC funded had the highest correlation with the projected impact of ending CDC funding for HIV testing (Spearman correlation coefficients: 0.94 and 0.71). The impact of cessation of CDC-funded testing was also negatively correlated (−0.58) with the urbanicity of states’ HIV epidemics: The more a state’s HIV epidemic was situated in rural areas, the greater the impact of removing CDC funding for HIV tests (Figure 4).

**Figure 4.**
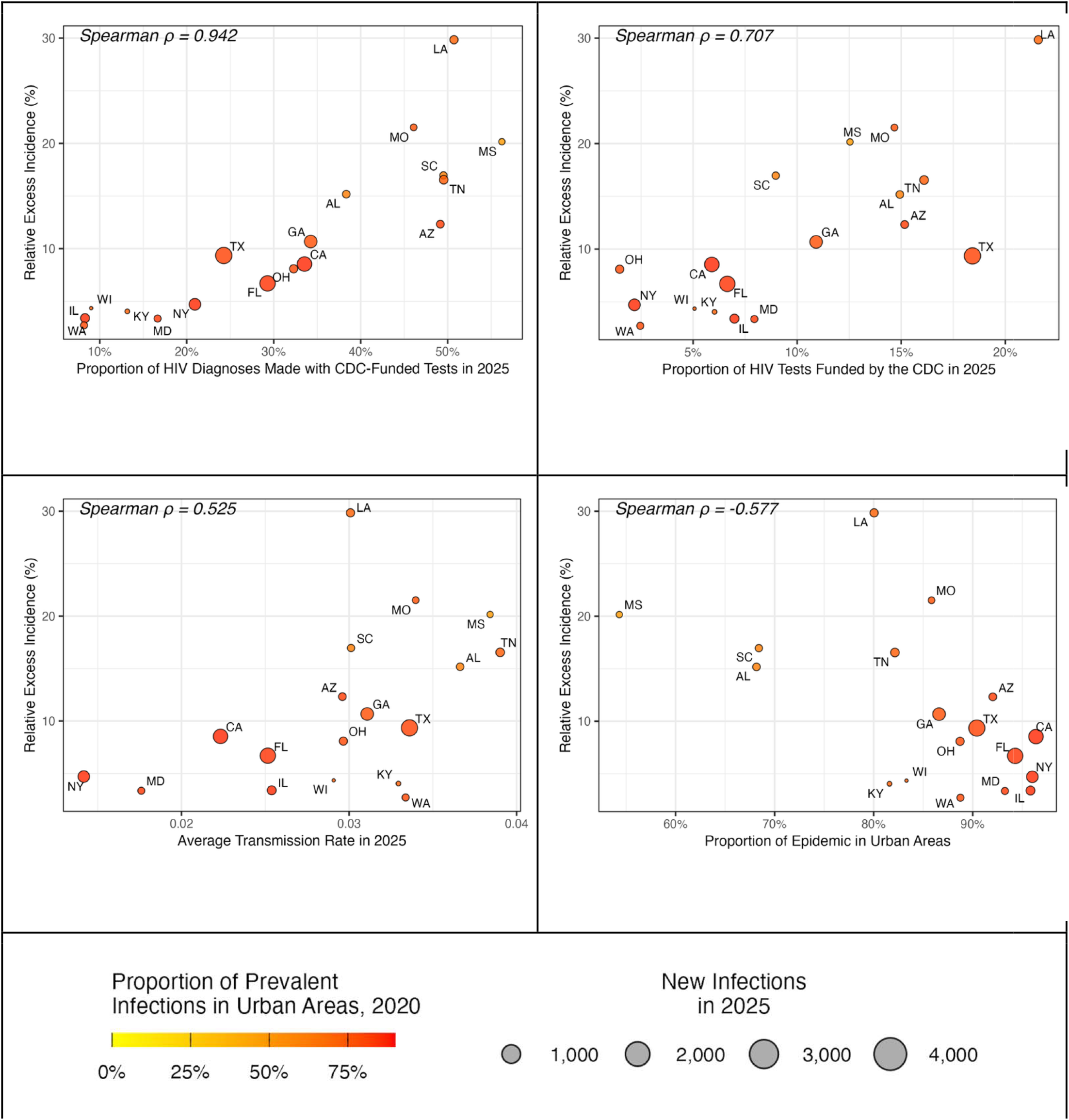
State-level Variation in Excess HIV Infections if CDC-funded HIV Testing Ends

Across all states, we project that 9.9 million HIV tests would be foregone from 2025 to 2030 in the “Cessation” scenario, yielding one excess infection for every 913 tests not funded by the CDC (95% CrI 453 to 2,145). The projected efficiency of CDC-funded tests varies between states, ranging from 137 (95% CrI 63 to 327) tests not done per one excess infection in Ohio to 4,396 (95% CrI 2,194 to 10,064) tests not done per one excess infection in Maryland (Supplement Figure S9).

The proportion of CDC-funded tests that would be done otherwise if CDC funding ends was strongly associated with the projected impact of funding cessation, with a partial rank correlation coefficient less than −0.99 in all states (Supplement Figure S10). In simulations where the lowest quintile (11 to 36% of CDC-funded tests) would still be performed in the absence of CDC funding, we project 19,145 excess cases between 2025 and 2030 (95% CrI 17,618 to 20,455). Conversely, in simulations where 63 to 88% of tests would still be performed, 6,620 (95% CrI 5,456 to7,925) excess infections are projected to occur if all CDC-funded HIV testing ends (Figure 5; Supplement Figures S11-13).

**Figure 5.**
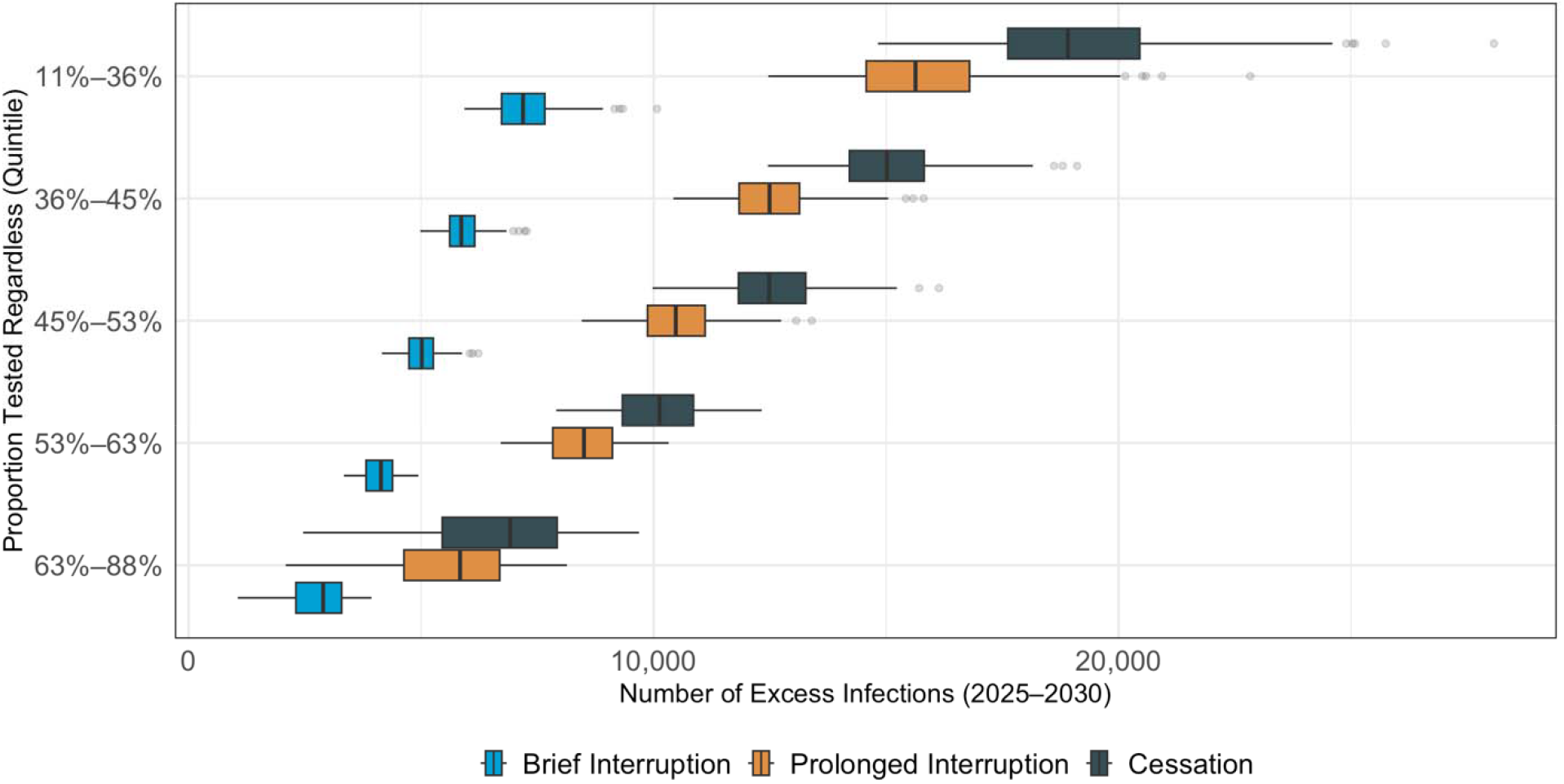
Excess HIV Infections 2025-2030 if CDC Funding for HIV Tests Ends, According to the Proportion of CDC-Funded Tests Performed Without CDC Funding

## Discussion

We used a mathematical HIV transmission model to estimate the impact of disruptions to CDC funding for HIV tests in 18 US states. Across all states, we project that complete cessation of CDC-funded HIV testing would lead to more than 12,000 additional HIV infections from 2025 to 2030 – a 10% increase vs. if testing continues uninterrupted. The excess would accrue over time, reaching 15% more infections by 2035. Even temporary interruptions to testing would also result in substantially more infections by 2030: an 8% increase if testing resumes in 2029 and a 4% increase if testing resumes in 2027. These projections assume there is some level (on average 50% across simulations) of replacement testing within the community; incidence would likely increase substantially if alternative access to testing did not materialize. The projected percentage of excess infections varied substantially between states, reaching as high as 30% by 2030 (9 to 60%) in Louisiana, the state with the highest proportion of HIV tests funded by the CDC.

Differences between states were most closely correlated with the number of tests funded by the CDC and the proportion of a state’s diagnoses that were made with CDC-funded tests. The impact of ending CDC funding for HIV tests was also correlated with the urban/rural distribution of HIV in the state: states with a more rural epidemic tended to have higher projected increases in incidence from disruptions to CDC-funded testing.

Our projections depended on the degree to which HIV tests would still be performed if CDC-funded tests become unavailable. If less than 36% of currently-CDC-funded tests are still performed in the absence of CDC funding, we project 19,145 excess infections from cessation of CDC-funded HIV testing versus 6,620 if more than 63% of such tests are still performed. The true proportion of CDC-funded tests that would be performed in the absence of CDC funding is unknown; there are no studies to our knowledge on the effects of widespread reductions in publicly funded HIV testing. We incorporated this uncertainty into our analysis by sampling a range of possible values across the 1,000 simulations in each state. Notably, it may be that cessation of public testing results in more efficient testing (for example, if people continue to prioritize tests that are more likely to be positive) or less efficient testing (if people at highest risk are also those least likely to still get tested).We therefore sampled a broad range around a mean of 50% of CDC tests being performed regardless: 95% of simulations had a value from 20 to 80%. Fundamentally, this quantity is uncertain, and our analysis reflects this uncertainty in broad credible intervals around our projections. The importance of this parameter implies that, if CDC funding for HIV tests does end, efforts to mobilize access to other means of HIV testing will be critical to mitigate the impact on local HIV epidemics.

There are few other published estimates of the impact of the CDC’s HIV testing activities. Hutchinson, et al., used a transmission model to estimate that CDC-funded HIV tests averted 3,381 new infections nationwide from 2007 to 2009, during which 2.8 million people were tested (824 people tested per infection averted)^19^. This is lower than the 12,719 infections averted over five years in 18 states that we project, although similar to our aggregate 913 tests per infection averted. Our model differs from Hutchinson, et al., in that it simulates transmission dynamics over time, such that an averted infection can also avert other infections through subsequent averted transmission. More recently, the CDC estimated that HIV prevention programs (including both testing and other prevention programs) prevented 9,000 infections between 2017 and 2021 - again lower than our estimate, in part due to the lack of incorporation of a dynamic modeling approach^20^.

As with any modeling study, our approach has several limitations. First, we focused on incident infections only; however, delayed diagnosis of HIV can also lead to increased morbidity and mortality. Second, we only model 18 US states. While the majority of HIV diagnoses in 2024 were made in these states, they may not reflect the full heterogeneity of HIV epidemics across the US. Third, in our interruption scenarios, we assume that CDC testing activities would return to their previous levels within a year, but it is possible that programs might recover more slowly. Finally, our projections assume no concurrent changes to HIV prevention and control efforts in the US. This is unlikely to be true; other disruptions to prevention activities would likely accompany cessation of CDC-funded testing, and future changes to Medicaid coverage may impact HIV screening and treatment.

Our approach also has several strengths. Using state-level models allows us to capture local-level heterogeneity in HIV epidemics and the particular ways they interact with testing funded by the CDC. Our Bayesian calibration process enables us to robustly recapitulate historical trends and characterize uncertainty in future projections. Finally, our projections are all available in an interactive web tool at jheem.org/cdc-testing, allowing local decision makers to consider the potential impact of changes to CDC testing programs in their setting.

In summary, using an HIV transmission model in 18 states, we project that even brief interruptions to CDC-funded HIV testing could lead to more than 5,000 excess HIV infections by 2030. Complete cessation of testing could lead to more than 13,000 additional infections over this time frame. These effects varied across states, with states that use more CDC-funded tests and states with more rural epidemics expected to see greater increases in transmission. These findings demonstrate the importance of maintaining CDC-funded testing activities in curbing the spread of HIV in the US.

Sample projections for Illinois, Texas, and Louisiana. Y-axes give the projected number of infections. Lines denote the mean across 1,000 simulations; ribbons give the 95% CrI. Green represents uninterrupted “Continuation” of CDC funding for HIV testing. In the other scenarios, funding stops in October 2025. In the “Cessation” scenario (navy blue), CDC-funded tests never resume. In “Prolonged Interruption” (orange), CDC-funded tests return to prior levels from January to December 2029. In “Brief Interruption” (light blue), testing recovers from January to December 2027. States chosen to represent three states across the spectrum of relative excess incidences. Times of reintroduction of testing (2027 and 2029) are shown as vertical dashed red lines.

The “Continuation” column gives the mean and 95% CrI, across 1,000 simulations, for projected incident HIV infections from 2025-2030 if CDC funding for HIV tests continues uninterrupted. The columns labeled “Number of Excess Infections” give the mean and 95% interval of the absolute number of excess HIV infections expected from 2025-2030 under three scenarios where funding is stopped in October 2025: “Cessation” (funding does not resume), “Prolonged Interruption” (testing returns to prior levels from January to December 2029), and “Brief Interruption” (testing recovers from January to December 2027). The columns labeled “Relative Excess Infections” give the percent change in projected incident infections, relative to “Continuation”. Cells are shaded according to the relative excess infections.

Boxplots display the projected percentage increase in new infections under three scenarios in which CDC funding for HIV testing ends in October 2025: “Cessation” (navy blue) – funding does not resume; “Prolonged Interruption” (orange) – testing returns to prior levels from January to December 2029; “Brief Interruption” (light blue) – testing recovers from January to December 2027. The value along the x-axis represents the relative increase in cases vs. a scenario where CDC-funded HIV tests continue uninterrupted. The dark vertical lines indicate the mean projection across 1,000 simulations, the boxes indicate interquartile ranges (IQR), and whiskers cover the 95% CrI. *The CrI has been truncated at 40%.

Each circle represents one state. The y-axis represents the average relative increase in projected HIV infections from 2025 to 2030 if CDC funding for HIV testing ends in October 2025 versus continuation at current levels, averaged across 1,000 simulations. The x-axis represents the average proportion of 2025 diagnoses that were made with CDC-funded tests in the state (Panel A), the average proportion of all tests in 2025 that were funded by the CDC (Panel B), the average transmission rate in 2025 (Panel C), or the proportion of prevalent HIV cases in 2021 that fell into rural areas with the state (Panel D). The size of the circle is proportional to the number of projected new diagnoses in 2025. Cities are shaded according to the proportion of prevalent cases that fall into urban areas. Correlation denotes the Spearman rank correlation.

Boxplots display the projected percentage increase in new infections under stratified by the simulated proportion of people who would be tested regardless of whether CDC-funded tests are available. Colors denote: “Cessation” (navy blue) – funding does not resume; “Prolonged Interruption” (orange) – testing returns to prior levels from January to December 2029; “Brief Interruption” (light blue) – testing recovers from January to December 2027. The value along the x-axis represents the relative increase in cases vs. a scenario where CDC-funded HIV tests continue uninterrupted. The dark vertical lines indicate the mean projection across 1,000 simulations, the boxes indicate interquartile ranges (IQR), and whiskers cover the 95% CrI.

## Data Availability

All code to reproduce this analysis can be found at https://github.com/tfojo1/jheem_analyses/.

## Acknowledgements

RB and ATF conceived of the study, performed primary analysis and initial drafting of the manuscript. MS, RF, KAG,MS and PK contributed to data collection and model development. All authors reviewed and edited drafts of the manuscript. All authors have no conflicts of interest to declare. This work is supported by the National Institutes of Health [grant number R01MD018539 to ATF supporting MS, PK, MS, DWD, DSB, RF]. All code to reproduce this analysis can be found at https://github.com/tfojo1/jheem_analyses/.

